# Oral Metformin May Exacerbate Diabetic Neuropathy: A Two-Sample Mendelian Randomization Study

**DOI:** 10.1101/2025.02.11.25322054

**Authors:** Xiaoyong Wang, Yujuan Wu, Ningning Ma, Jueying Chen, Yuanjin Chen, Jiaoling Shi

## Abstract

**Objective:** Given the important role of metformin in diabetes treatment and the uncertainty regarding its relationship with diabetic neuropathy, this study aims to assess the causal relationship between metformin and diabetic neuropathy using Mendelian randomization.

**Methods:** Instrumental variables were derived from publicly available genome-wide association study datasets. Two study cohorts were assigned: a training cohort (exposure ID: ukb-a-159) and a testing cohort (ID: ukb-b-14609), with the outcome ID being finn-b-DM_NEUROPATHY for both.

The primary analysis used inverse variance weighting (IVW) for Mendelian randomization, supplemented by weighted median (WMed), weighted mode (WM), simple mode (SM), and MR Egger regression (MER). To test for heterogeneity, pleiotropy, and publication bias, we conducted leave-one-out sensitivity analyses, MR Egger regression, and further assessed with funnel plots.

**Results:** Mendelian randomization analysis showed a significant positive causal relationship between metformin and diabetic neuropathy in both the training set (IVW: logOR 14.73, 95% CI [9.60, 19.86], p=1.78E-08) and the testing set (IVW: logOR 15.83, 95% CI [11.10, 20.56], p=5.54E-11). The WMed, WM, SM, and MER models all consistently supported this conclusion, with no evidence of pleiotropy or heterogeneity, indicating robust results.

**Conclusion:** From an epidemiological perspective, this study reveals a significant positive correlation between metformin use and diabetic neuropathy through Mendelian randomization analysis. This finding not only provides new insights into the field of diabetes treatment but also suggests that the use of metformin may be associated with the potential side effect of increased risk of diabetic neuropathy.

## Background

Diabetic neuropathy is one of the most common chronic microvascular complications of diabetes, affecting cranial nerves, central nerves, and peripheral nerves, and resulting in symptoms such as numbness, pain, decreased sensation, and muscle atrophy in the distal limbs[1]. The onset of diabetic neuropathy is insidious and progresses slowly, yet it severely impacts patients’ quality of life. Its pathogenesis is closely related to metabolic disorders caused by hyperglycemia, damage to neurotrophic microvessels, and neurotrophic disorders[2]. Despite the availability of various treatment methods, the prevention and treatment of diabetic neuropathy remain suboptimal[3], necessitating new therapeutic strategies to slow disease progression.

Metformin, which was formally introduced into clinical practice in 1957, has become the first-line treatment for type 2 diabetes[4]. It not only effectively reduces blood glucose levels but also offers multiple benefits, including cardiovascular protection and improved insulin resistance[4]. Despite the broad benefits of metformin in diabetes treatment, its relationship with diabetic neuropathy remains controversial[5]. Some studies have suggested that metformin use may be associated with a reduced risk of diabetic neuropathy, potentially due to its effects on improving insulin resistance, lowering blood glucose levels, and exerting anti-inflammatory and antioxidant actions[6]. However, other studies have proposed that long-term use of metformin may have adverse effects on the nervous system, especially at high doses and in specific patient populations (e.g., patients under 60 years of age with fewer comorbidities)[8]. These conflicting results suggest that the relationship between metformin and diabetic neuropathy may be influenced by multiple factors, including genetic background, drug dosage[9], treatment duration[10], and overall patient health status[8].

Given the important role of metformin in diabetes treatment and the uncertainty regarding its relationship with diabetic neuropathy, this study aims to assess the causal relationship between metformin and diabetic neuropathy using Mendelian randomization (MR). By leveraging genetic variants as instrumental variables (IVs), this study seeks to minimize the impact of confounding factors and more accurately evaluate the effect of metformin on the risk of diabetic neuropathy. This will provide new evidence and guidance for the application of metformin in the prevention and treatment of diabetic neuropathy.

## 1 Methods and Materials

This study is a secondary data review of existing databases. MR is a method that uses genetic variants of exposure as instrumental variables (IVs) to identify causal relationships between exposure phenotypes and outcomes. It leverages publicly available datasets from large-scale genome-wide association studies (GWAS) for "exposure" and "outcome" and overcomes typical limitations of observational studies.

It is a statistical method based on whole-genome sequencing data, designed to reveal causal relationships. It is based on three core assumptions: 1) Relevance assumption (or correlation assumption): Genetic variants (single nucleotide polymorphisms, SNPs) must have a strong association with the exposure factor under study, ensuring that the selected genetic variants can serve as effective proxy variables for the exposure[11]. 2) Independence assumption: The instrumental variables (i.e., genetic variants) must be independent of any known or unknown confounding factors to avoid interference from confounders in the causal relationship, ensuring that the inferred causal relationship is accurate[12]. 3) Exclusivity assumption: Genetic variants can only affect the outcome through the exposure factor and not through other direct causal pathways, ensuring that the impact of genetic variants on the outcome is entirely mediated through the exposure factor, thereby accurately inferring the causal relationship between the exposure factor and the outcome[13].

### 1.1 Data Sources

In MR analyses, UKB and FinnGen are two frequently mentioned large-scale databases or cohort studies that provide valuable data resources for MR analysis[14]. UKB, the UK Biobank, is a large cohort study comprising hundreds of thousands of individuals[15]. It provides rich individual-level data, including genomic data, clinical data, and lifestyle data, which are of significant value for studying causal relationships of various diseases and traits. In MR analysis, data from UKB are often used to validate and analyze research results. Researchers can utilize the genomic data from UKB, combined with GWAS data of exposure factors and outcomes, to conduct MR analysis and assess the causal relationships between exposure factors and outcomes. The large scale of UKB data, along with its GWAS results for various diseases and traits, enables researchers to validate and analyze causal relationships in larger sample sizes, thereby enhancing the reliability and accuracy of the results.

FinnGen is a large-scale genome-wide association study cohort in Finland, aimed at identifying genetic variants associated with various diseases and traits through GWAS[16]. Similar to UKB, data from FinnGen can also be used for MR analysis. Researchers can use the genomic data and GWAS results from FinnGen, combined with data on exposure factors and outcomes, to evaluate causal relationships. FinnGen’s data, characterized by high homogeneity and detailed phenotypic information, offer advantages in identifying genetic variants associated with diseases. Additionally, FinnGen provides rich publicly available data, making it easier for researchers to access and use these data. The selection of appropriate databases or cohort studies is crucial in MR analysis. UKB and FinnGen, as two large-scale, high-quality databases, provide reliable data support for MR analysis.

In this study, we utilized two cohorts derived from the UK Biobank (UKB), focusing on patients using metformin as the exposure factor. The unique identifiers (IDs) of the two datasets were ukb-a-159 and ukb-b-14609, respectively. The data for the ukb-a-159 cohort originated from a study led by Professor Neale in 2017, which analyzed 10,894,596 single nucleotide polymorphisms (SNPs) based on over 337,000 samples, using the HG19/GRCh37 human genome build as the reference standard[17].

The other dataset, ukb-b-14609, was derived from a GWAS conducted by Ben Elsworth et al. in 2018, involving 454,805 samples and 9,851,867 SNPs, also using the HG19/GRCh37 human genome reference sequence[17]. Notably, both studies focused on European populations, ensuring the homogeneity and comparability of the data. In this study, the ukb-a-159 cohort was used as the training set to construct and validate our analysis model, while the ukb-b-14609 cohort served as the testing set to evaluate the model’s predictive performance.

Additionally, to investigate diabetic neuropathy as the outcome, we incorporated cohort data from the FinnGen database, with the ID finn-b-DM_NEUROPATHY. This was a binary classification study focused on diabetic neuropathy, involving 1,415 case individuals and 162,201 control individuals, analyzing 16,380,195 SNPs. The study also used the HG19/GRCh37 human genome reference sequence, ensuring consistency and comparability of the data[18].This study is a secondary data review utilizing existing databases. Therefore, no additional ethical approvals were required.

### 1.2 Selection of SNPs for Exposure and Outcome

Extracting high-quality genetic variables is a key step in Mendelian randomization studies to avoid interference from reverse causation and confounding factors. By setting reasonable and stringent significance thresholds and implementing clumping processes, the selected variables can have strong associations with the exposure factor while being relatively independent of each other, thereby enhancing the accuracy and reliability of Mendelian randomization studies[19]. For the exposure factor, a widely accepted significance threshold of p < 5×10^-8 was used, meaning that only SNPs with a p-value less than 5×10^-8 in GWAS would be selected as instrumental variables. This threshold ensures that the selected SNPs have sufficient statistical significance[20].

During the clumping process, a linkage disequilibrium (LD) threshold of r² = 0.001 was used, meaning that any two SNPs with an r² value greater than 0.001 would be considered highly correlated, and only one would be retained in the same clump. A physical distance threshold of kb = 10000 was applied, meaning that within a genomic distance of 10,000 kb (i.e., 10 Mb), only the SNP with the smallest p-value would be retained, while others would be removed as highly correlated.

### 1.3 Statistical Analysis

The two-sample MR analysis was conducted using R software (version 4.4.2) and the TwoSampleMR package (version 0.6.8). In Mendelian randomization analysis, the Inverse Variance Weighting (IVW) model was the primary analytical method.

Additionally, several other methods were employed to estimate causal effects, including Weighted Median (WMed), MR-Egger regression (MER), Simple Model (SM), and Weighted Model (WM). These methods were used as supplementary approaches to provide a more comprehensive assessment of causal effects.

The mr function from the TwoSampleMR package was used to perform Mendelian randomization analysis, identifying pairs with more than 10 eligible SNPs [20, 21]. The analysis calculated the effect estimates (b-values) of exposure on outcomes, their standard errors (SE), p-values, and log odds ratios (Log(OR)) with confidence intervals. The mr_egger_regression function was used to test for heterogeneity and assess pleiotropy through intercept analysis. If pleiotropy was detected, the interpretation of results would become complex, making it difficult to determine whether the outcome (diabetic neuropathy) was caused by the exposure (metformin), rendering the study inconclusive. If the heterogeneity test showed p < 0.05, a random-effects model was used in subsequent analyses; otherwise, a fixed-effects model was applied.

Leave-One-Out (LOO) sensitivity analysis was conducted to evaluate the stability of the Mendelian randomization results by sequentially excluding instrumental variables. Publication bias was further assessed using funnel plots. The F-statistic was calculated to estimate weak instrument bias, with F < 10 indicating potential bias [22]. The analysis results were exported to Excel and PDF files, including forest plots, pleiotropy plots, sensitivity analysis plots, and funnel plots.

## 2 Results

### 2.1 Training Set Analysis: The Relationship Between Metformin and Diabetic Neuropathy

After this step, a total of 45 SNPs were retained for subsequent analysis (see Supplementary Material 1).

The MRE regression heterogeneity test results showed significant heterogeneity in our data (Q=120.1205042, Q_pval=3.23E-09) (see Supplementary Material 2). To further understand this heterogeneity, we drew and analyzed single SNP forest plots (see Figure 1-1-A), leave-one-out sensitivity test forest plots (see Figure 1-1-B), and funnel plots (see Figure 1-1-C). In these detailed analyses, we found that the results of the target SNP rs9273268 were significantly deviated (pval.outcome=4.00313E-21). It should be noted that rs9273268 was included in this analysis through its proxy SNP rs17612496 (see Supplementary Material 3). Given that the significant deviation of rs9273268 may have an adverse effect on the overall results, we cautiously decided to exclude it from subsequent analyses. Therefore, the final number of SNPs used for analysis was 44 (see Supplementary Material 4).

**Figure 1-1-A:**
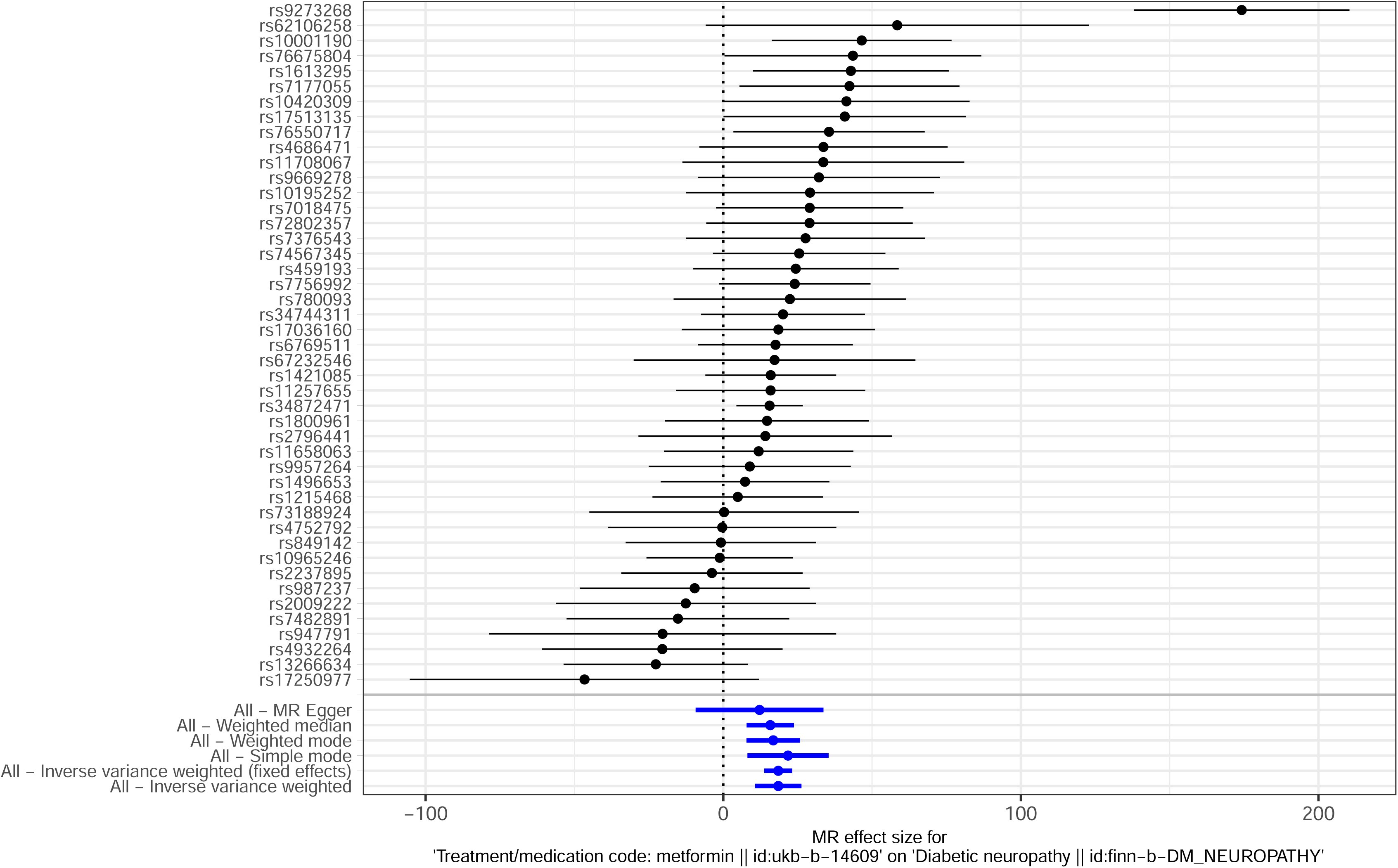
Forest plot of single SNPs in the training set analysis before excluding rs9273268.

**Figure 1-1-B:**
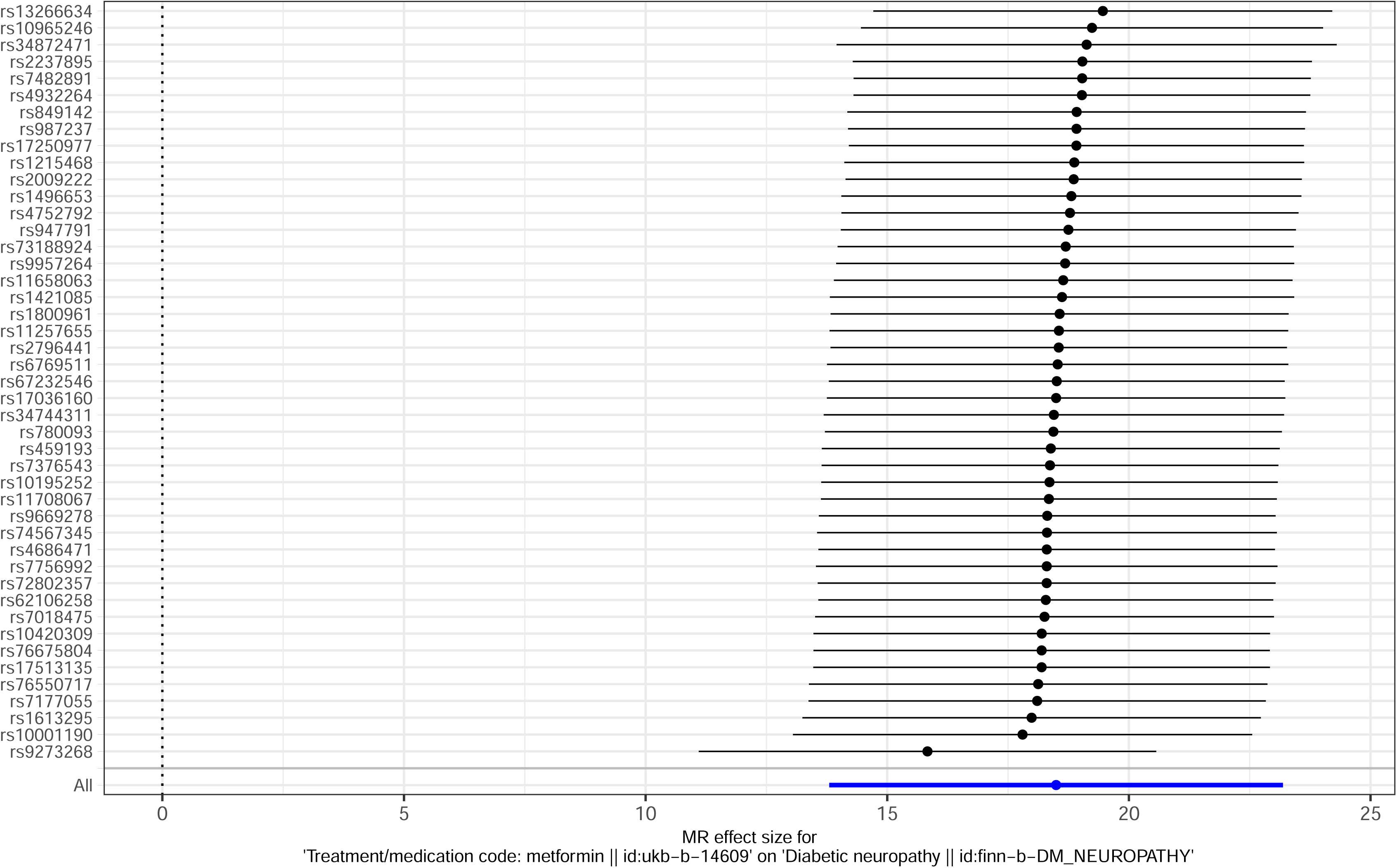
Leave-one-out sensitivity test forest plot in the training set analysis before excluding rs9273268.

**Figure 1-1-C:**
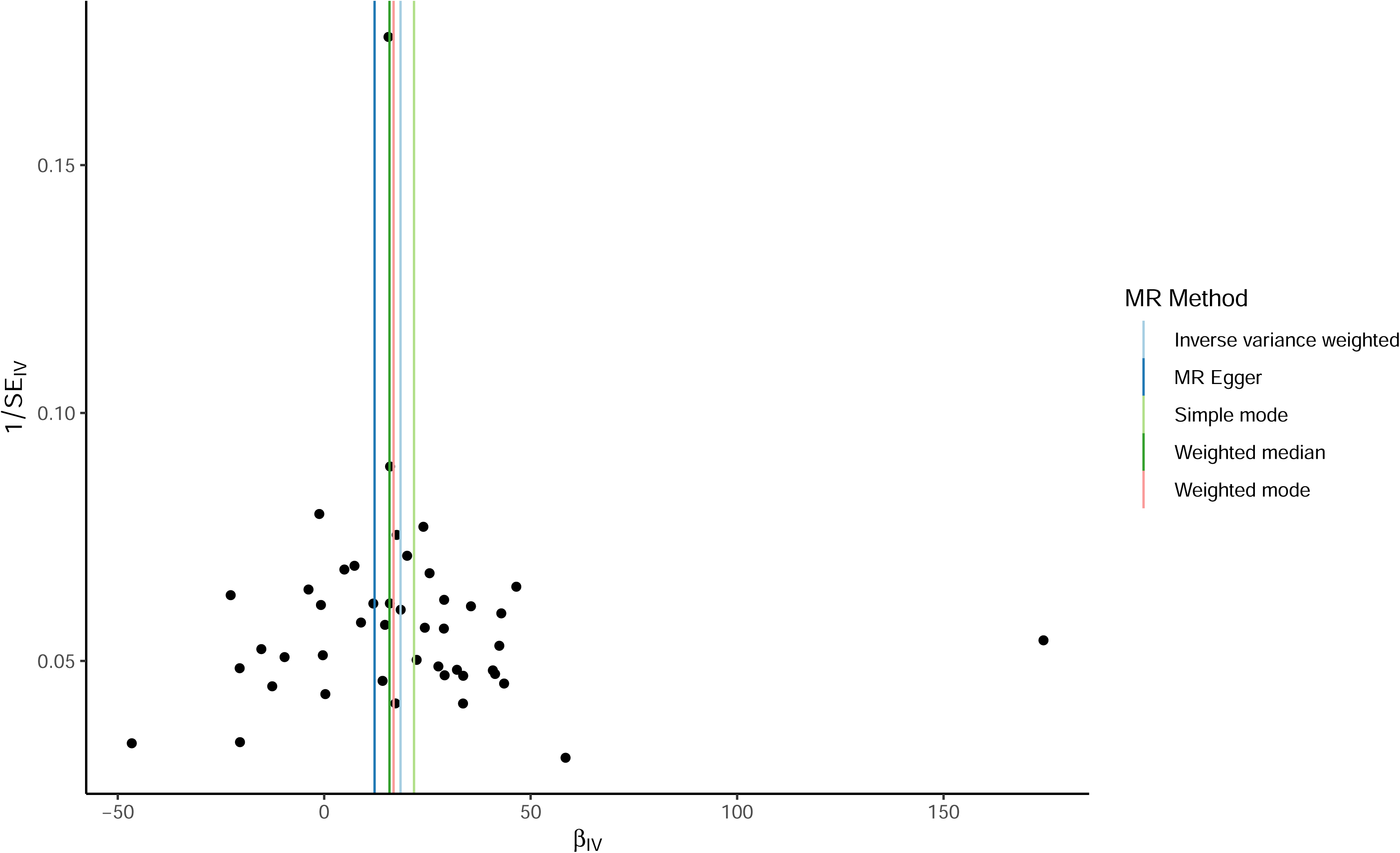
Funnel plot in the training set analysis before excluding rs9273268.

After excluding rs9273268, we performed MRE regression analysis again, and the results did not show significant heterogeneity (Q=48.87, Q_pval=0.22) (see Supplementary Material 5). Based on this result, we used the fixed-effect model in the IVW analysis. The IVW analysis results also revealed a significant positive causal relationship between metformin and diabetic neuropathy (logOR=15.83, 95% CI [11.10, 20.56], p=5.54E-11). In addition, WMed analysis (OR=15.80, 95% CI [7.68, 23.91], p=1.36E-04), WM analysis (logOR=16.76, 95% CI [7.64, 25.89], p=8.17E-04), SM analysis (logOR=21.48, 95% CI [7.62, 35.33], p=4.04E-03), and MER analysis (logOR=15.62, 95% CI [1.73, 29.50], p=3.30E-02) all consistently supported a significant positive causal relationship between metformin and diabetic neuropathy (see Figure 1-2). LOO sensitivity test further confirmed the robustness of these results (see Figure 1-2-A). In addition, the results of MR-Egger regression analysis

**Figure 1-2:**
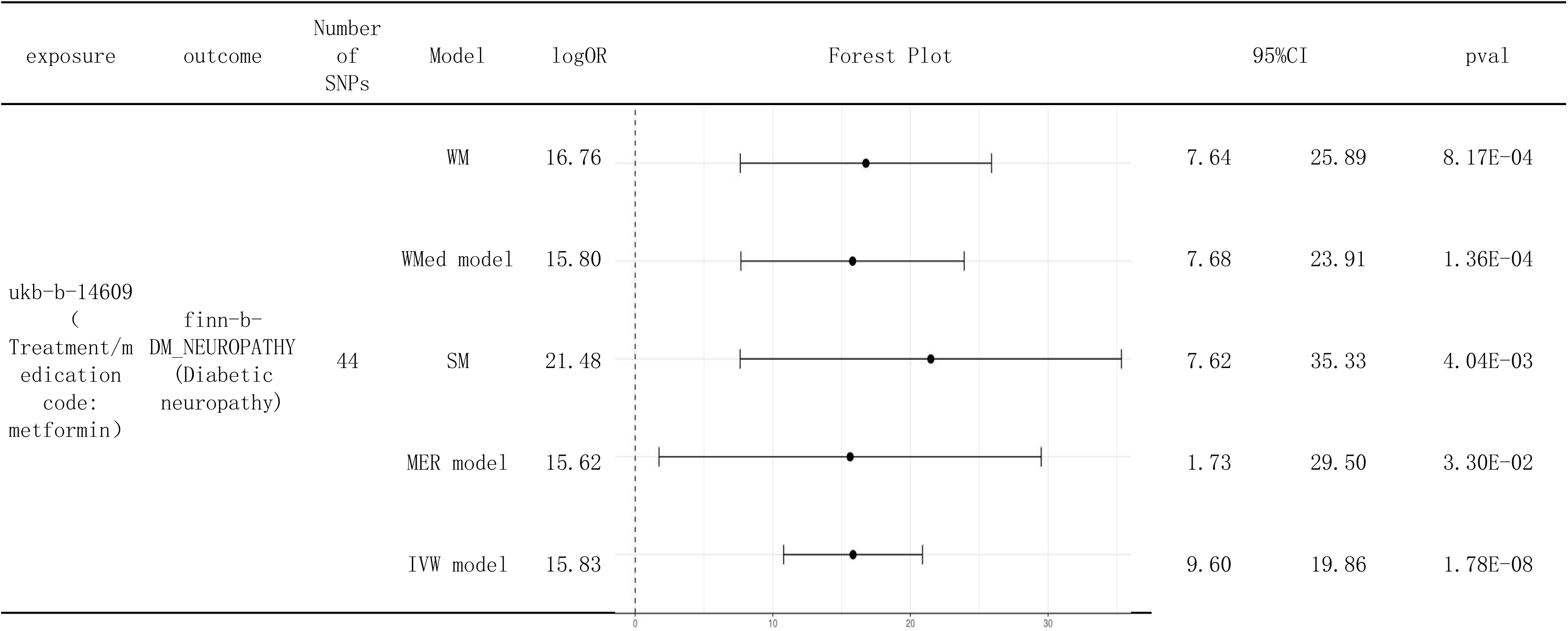
Forest plots of five analysis models (IVW, WMed, WM, SM, MER) in the training set after excluding rs9273268

**Figure 1-2-A:**
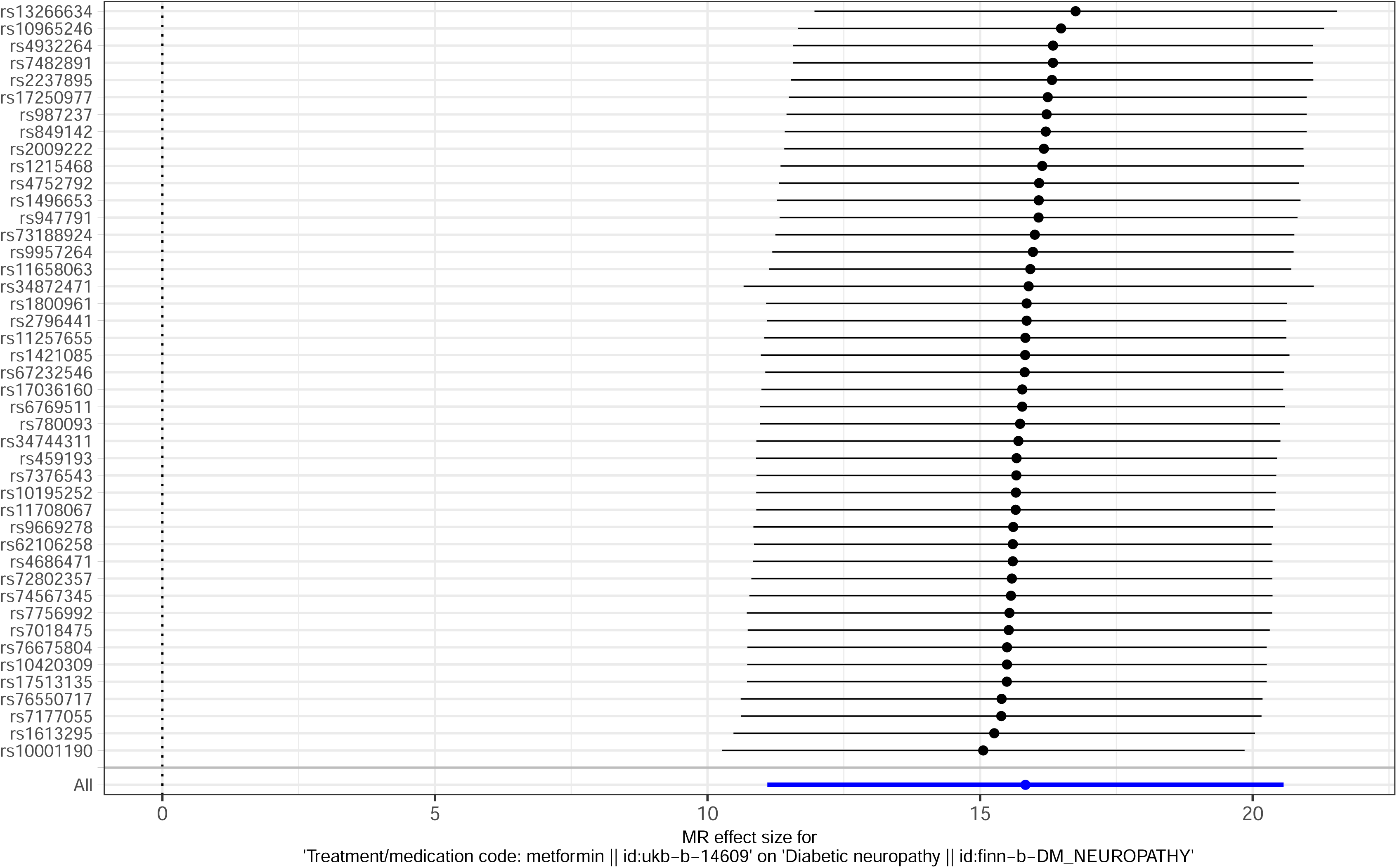
Leave-one-out sensitivity test forest plot in the training set analysis after excluding rs9273268.

### 2.2 Test Set Analysis: The Relationship Between Metformin and Diabetic Neuropathy

In the test set MR study, 26 SNPs were identified in common between the exposure (ID: ukb-a-159) and the outcome. Two SNPs (rs649698 and rs649698) were removed due to being palindromic with intermediate allele frequencies, leaving 24 SNPs for analysis (see Supplementary Material 6). The MR Egger regression test showed no evidence of significant heterogeneity (Q=27.01, Q_pval=0.26) (see Supplementary Material 7), so the fixed-effect model was used in the IVW analysis. The IVW analysis found a significant positive causal relationship between metformin and diabetic neuropathy (logOR=14.73, 95% CI [9.60, 19.86], p=1.78E-08), which was supported by WMed (OR=14.92, 95% CI [7.32, 22.52], p=1.19E-04), WM (logOR=14.98, 95% CI [6.48, 23.47], p=2.16E-03), SM (logOR=15.41, 95% CI [3.01, 27.80], p=2.30E-02), and MER (logOR=14.29, 95% CI [0.75, 27.82], p=5.05E-02) analyses (see Figure 2-1). LOO sensitivity test confirmed the robustness of the results (see Figure 2-2-A). In the MR-Egger regression (intercept=4.25E-04, se=0.02, p=0.99), no pleiotropy was found (see Figure 2-2-B).

**Figure 1-2-B:**
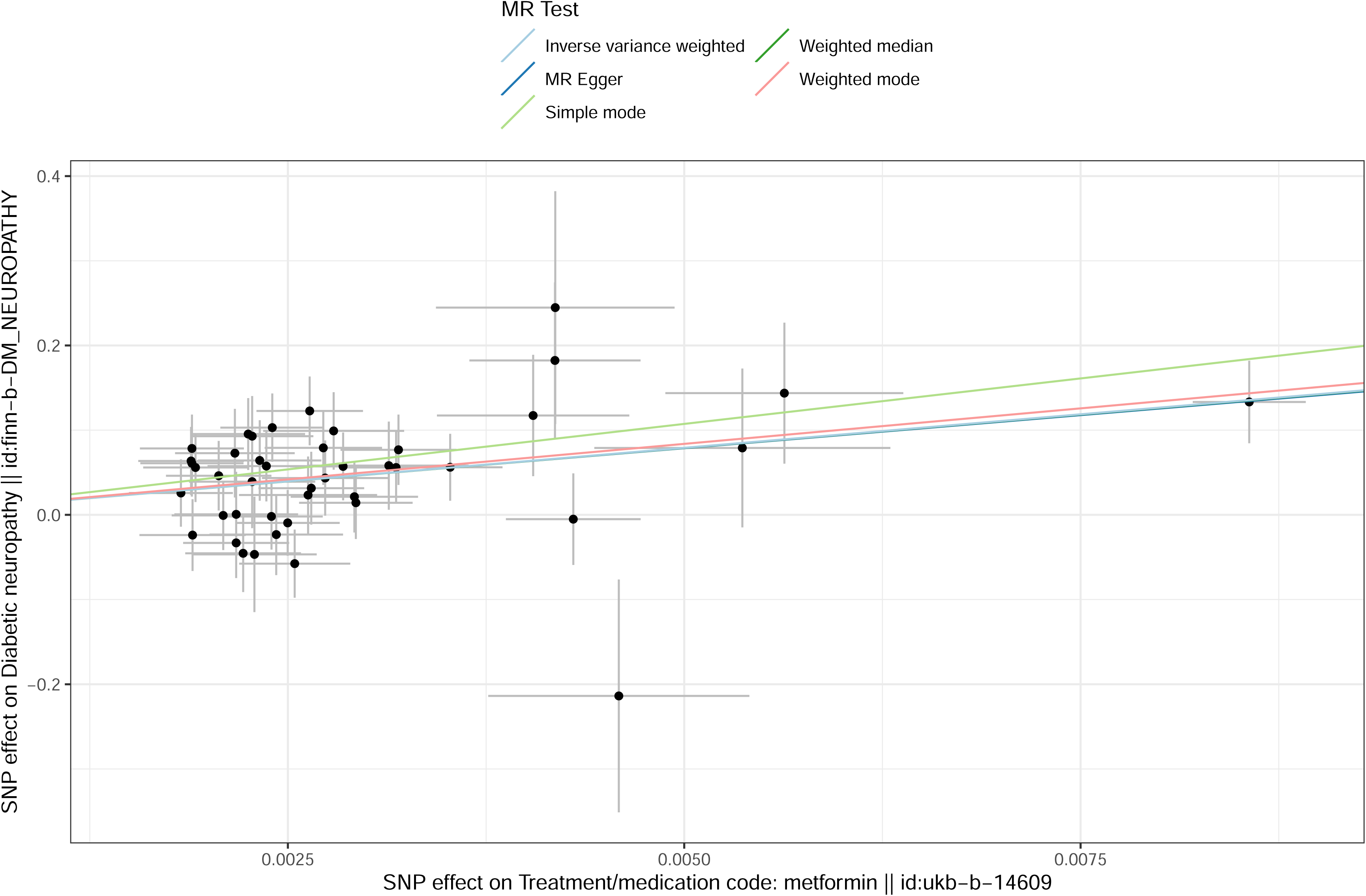
Scatter plot of pleiotropy in the training set after excluding rs9273268.

**Figure 2-1:**
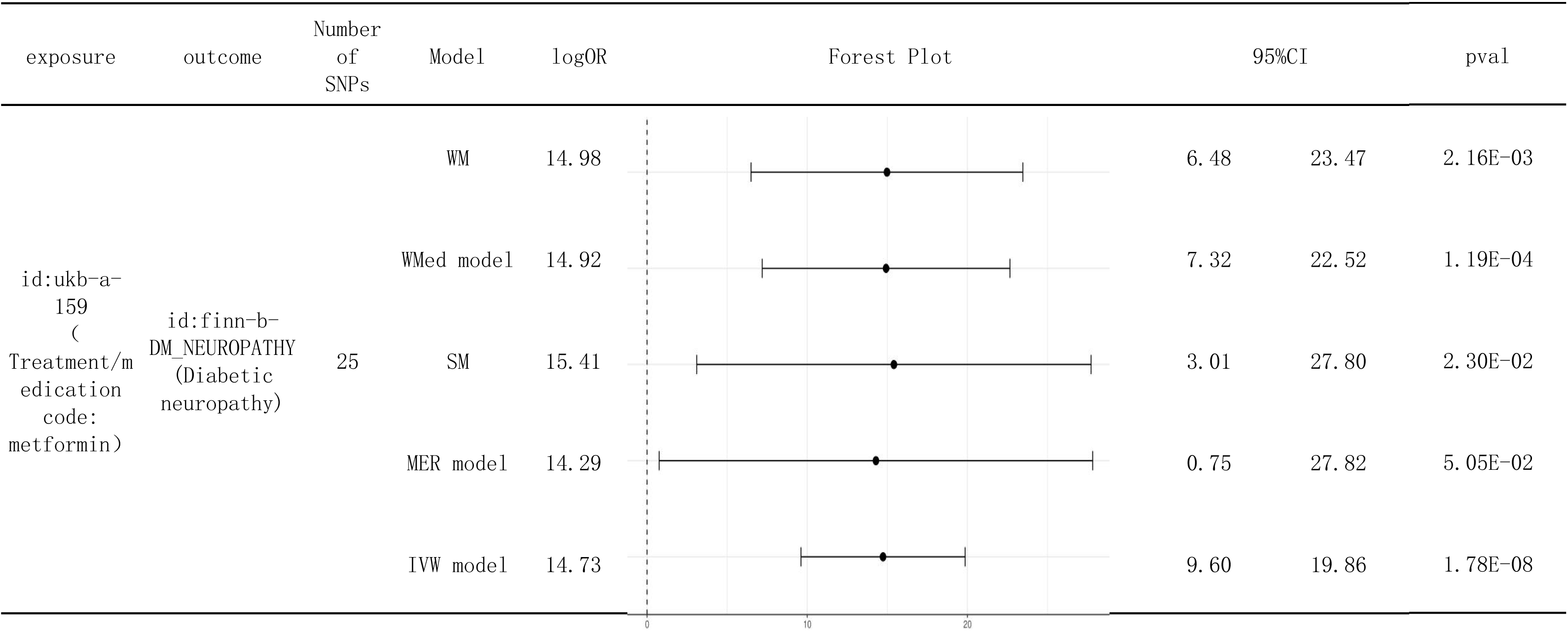
Forest plots of five analysis models (IVW, WMed, WM, SM, MER) in the test set.

**Figure 2-2-A:**
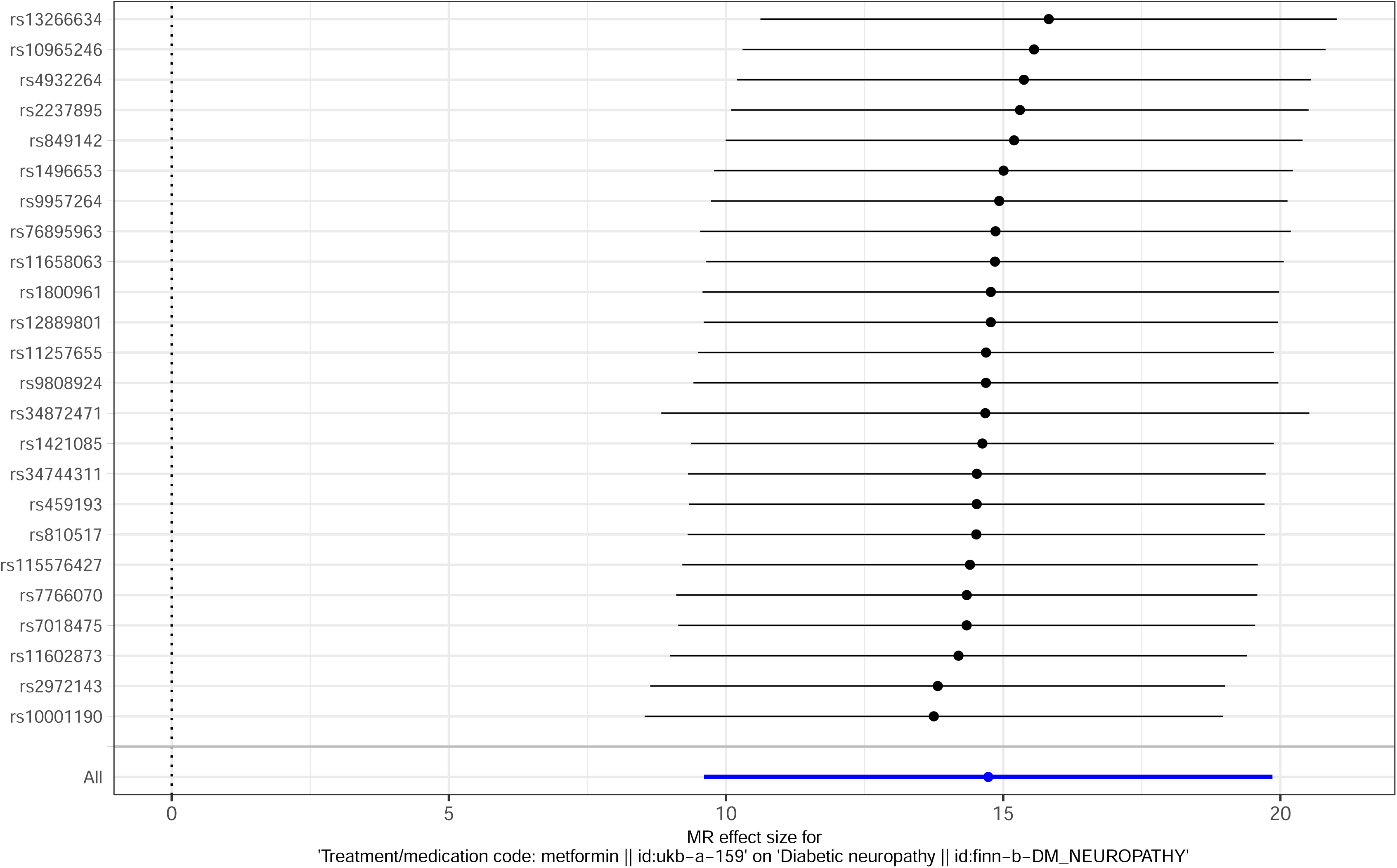
Leave-one-out sensitivity test forest plot in the test set.

**Figure 2-2-B:**
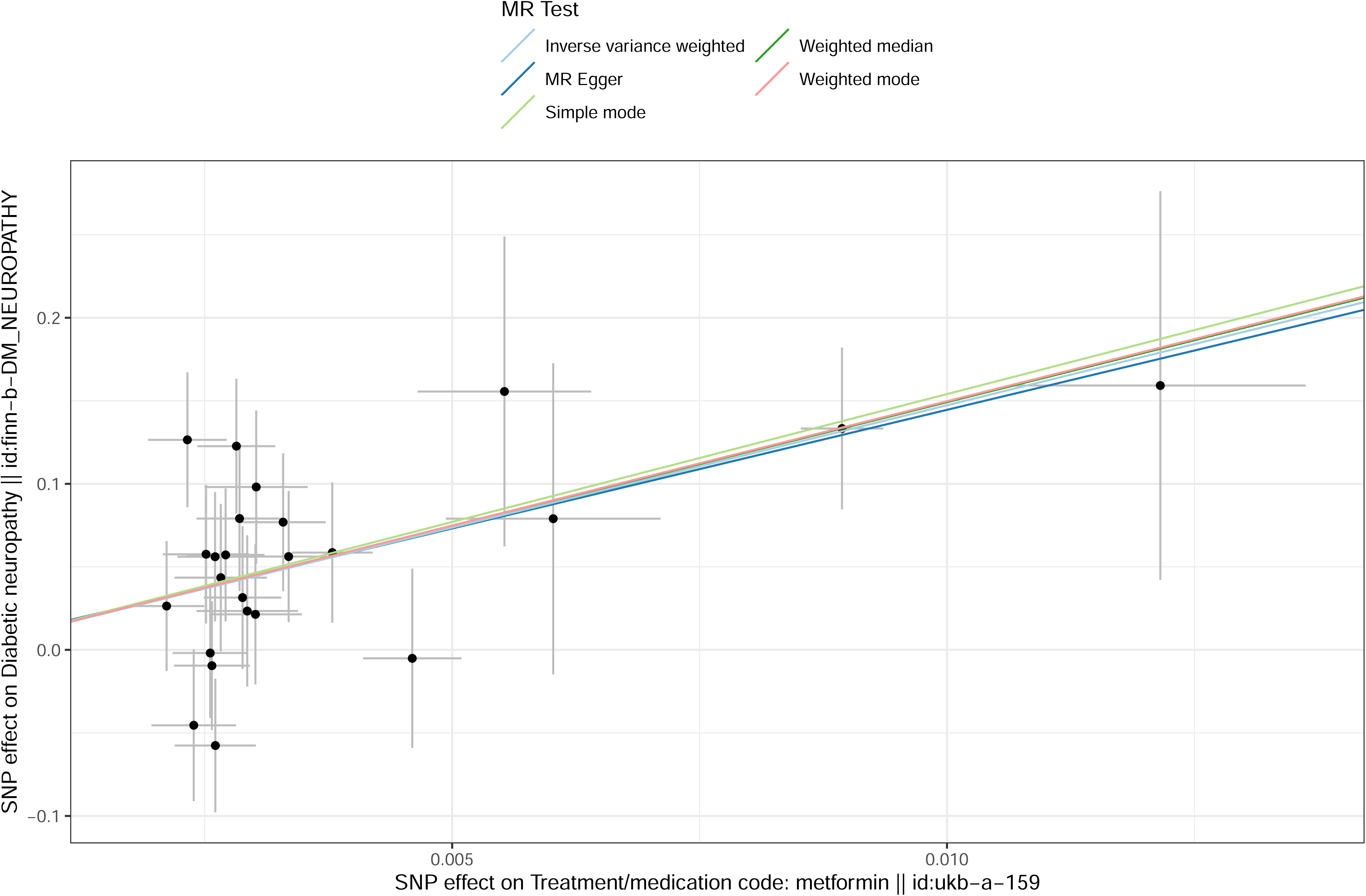
Scatter plot of pleiotropy in the test set.

## 3 Discussion

Mendelian randomization (MR) has gained widespread attention and application in public health and medicine in recent years as an innovative epidemiological method. By leveraging genetic variants as instrumental variables, MR explores the potential causal relationships between diseases and exposures. Compared to traditional observational studies, MR effectively avoids common issues such as confounding bias and reverse causation, significantly enhancing the reliability and accuracy of findings. This is because genetic variants are randomly distributed in the population, free from the interference of external confounding factors, thus simulating a naturally occurring randomized controlled trial.

In this study, we used the MR method to investigate the causal relationship between metformin and diabetic neuropathy. The IVW analysis in the training set strongly supported a significant positive causal relationship between metformin and diabetic neuropathy. This conclusion was further validated in multiple analytical models, including WMed, WM, SM, and MER, demonstrating high robustness. LOO sensitivity analysis also confirmed the reliability of these findings. Additionally, MR-Egger regression showed no evidence of pleiotropy or heterogeneity, further strengthening our confidence in the conclusion.

In the testing set, we observed a significant positive association between metformin and diabetic neuropathy, which remained consistent and stable across various analytical methods and sensitivity tests. Although metformin is a standard treatment for type 2 diabetes, with significant clinical value in controlling blood glucose levels and potentially delaying neuropathy progression, our results suggest that metformin use may also increase the risk of diabetic neuropathy.

Our findings are consistent with previous cohort studies. According to Serra MC et al., compared to patients treated with metformin for at least 6 months but less than 18 months, veterans aged 50 years or older treated with metformin for at least 18 months had a significantly higher risk of diabetic peripheral neuropathy (DPN), approximately 2-3 times higher [10]. Hashem et al. found in a prospective clinical study that metformin use was associated with exacerbated DPN, especially within the first 6 months of treatment and more prominently in high-dose groups [9]. Hussain et al. [23] further explored this phenomenon, suggesting that metformin may lead to vitamin B12 (cobalamin) deficiency by interfering with calcium-dependent membrane activity and altering gut microbiota. Vitamin B12 deficiency can subsequently cause progressive demyelination, peripheral neuropathy, and hematological abnormalities. Moore et al. [7] also noted that metformin use may be associated with cognitive impairment. These findings highlight the importance of routine monitoring, early detection, and personalized intervention measures when using metformin to treat diabetes.

From an epidemiological perspective, our results suggest a significant link between metformin use and an increased risk of diabetic neuropathy. Biologically, this conclusion may be closely related to vitamin B12 deficiency caused by metformin use. Therefore, we recommend closely monitoring vitamin B12 levels in patients using metformin. Patients with borderline vitamin B12 levels should regularly test for methylmalonic acid and homocysteine to detect vitamin B12 deficiency early and take appropriate measures, including timely supplementation. However, we cannot rule out the potential role of other biological mechanisms in the association between metformin and diabetic neuropathy, and further in-depth research is needed.

Despite the strengths in design and methodology, our study has some limitations. For example, the selection of SNPs and the use of proxy SNPs may be influenced by factors such as linkage disequilibrium, sample size, and population differences. Future studies should consider these factors and explore more precise and comprehensive methods to assess the relationship between metformin and diabetic neuropathy, providing more reliable guidance for clinical practice.

## 4 Conclusion

Using the innovative epidemiological method of Mendelian randomization, this study explored the potential causal relationship between metformin and diabetic neuropathy. Through rigorous data analysis and validation using multiple analytical models, we concluded that there is a significant positive association between metformin use and diabetic neuropathy. This finding not only provides a new perspective on the role of metformin in diabetes treatment but also highlights a potential adverse effect of metformin, specifically the increased risk of diabetic neuropathy. Future research should further investigate the underlying mechanisms of this relationship and assess the risks and benefits of metformin use in different patient populations.

## Data Availability

All data produced in the present study are available upon reasonable request to the authors
All data produced in the present work are contained in the manuscript
All data produced are available online at：https://gwas.mrcieu.ac.uk/

## Acknowledgements

We sincerely thank all the participants who generously provided their data, and the researchers and scientists for their commitment to collecting and sharing summary-level data from GWAS.

## Authors’ contributions

Data curation, Yuanjin Chen, Jiaoling Shi and Jueying Chen; formal analysis, Ningning Ma; project management, Xiaoyong Wang and Yujuan Wu; writing - original draft, Xiaoyong Wang.

## Data availability

All data generated or analysed in this study are included in the published article [and its Supplementary Information files].

## Contributor information

Xiaoyong Wang

*Corresponding author: Yujuan Wu

